# Invasive Brain Mapping Identifies Personalized Therapeutic Neuromodulation Targets for Obsessive-Compulsive Disorder

**DOI:** 10.1101/2025.03.14.25323348

**Authors:** A Moses Lee, Audrey Kist, John Alvarez, Kristin K Sellers, Ankit N Khambhati, Leo P Sugrue, Lee B Reid, Kelly Kadlec, Joline M Fan, Anusha B Allawala, Caroline A Racine, Tenzin Norbu, Dani Astudillo, Alexandra G Tremblay-McGaw, Natalie Becker, Ahmad Alhourani, Philip A Starr, Edward F Chang, Andrew D Krystal

**Author notes:** Correspondence should be addressed to: A Moses Lee, MD PhD, Nancy Friend Pritzker Psychiatry Building, 675 18^th^ Street, San Francisco, California 94107. These authors contributed equally (Edward F Chang, Andrew D Krystal).

## Abstract

Deep brain stimulation has been used to treat severe, refractory obsessive-compulsive disorder (OCD) with variable outcomes across multiple anatomical targets. To overcome these limitations, we developed an invasive brain mapping paradigm in which electrodes were implanted across the OCD cortico-striato-thalamo-cortical circuit in a single individual. We then performed extensive stimulation mapping during a multi-day inpatient stay to identify personalized therapeutic targets and characterize their downstream circuit effects. We found two targets within the right ventral capsule (VC) that acutely reduced OCD symptoms. Prolonged VC stimulation suppressed high frequency activity within the structurally and functionally connected orbitofrontal cortex, which encoded the severity of OCD symptoms. These VC sites were implanted for DBS and combined stimulation of these targets led to a rapid therapeutic response. This case provides the first proof-of-concept that invasive brain mapping can be used to guide a novel personalized, multi-site neuromodulation approach to treat refractory OCD.

## Introduction

Deep brain stimulation (DBS) is a treatment for severe, refractory obsessive-compulsive disorder (OCD). Multiple DBS targets, such as the ventral anterior limb of the internal capsule, ventral striatum, bed nucleus of the stria terminalis, and subthalamic nuclei, have been used to treat OCD with response rates of ∼60% across sites [1, 2]. The lack of consistency in the response is hypothesized to be due to factors such as heterogeneity in circuit abnormalities underlying the disorder, anatomical variation across individuals, and lack of target engagement. The variable response to DBS delivered to each of these sites underscores the need to develop an approaches that can identify individualized therapeutic targets to optimize outcomes for OCD.

To address these challenges, we developed a protocol that aimed to identify personalized neuromodulation targets. Adapting methods commonly used in epilepsy to inform surgical treatment of epilepsy, our protocol involves the implantation temporary intracranial electrodes across the cortico-striato-thalamo-cortical (CSTC) circuits in a set of evidence-based candidate targets of neuromodulation for OCD. Our clinical goal was to perform extensive stimulation mapping to identify therapeutic sites that acutely reduced OCD symptoms. The invasive brain monitoring stage also provided us with a rare opportunity to identify personalized electrophysiological biomarkers of OCD symptom within the CSTC circuit and verify that acutely therapeutic stimulation modulated these biomarkers of target engagement using a multi-modal approach combining intracranial recordings and imaging. We then aimed to implant the top personalized candidate targets to implement a novel combined multi-site neuromodulation protocol that was aimed at achieving greater control of symptoms.

Here, we present results from our first patient to undergo our protocol. The patient was a woman in her 20s with extreme, treatment-refractory OCD (Yale Brown Obsessive-Compulsive Scale Y-BOCS: 36). Her OCD symptoms mostly consisted of harm-based obsessions associated with checking compulsions. These symptoms were disabling, occupied most of her waking hours, caused her to leave nursing school, and prevented her from driving or leaving home. She had not responded to multiple trials of serotonin reuptake inhibitors, augmentation strategies, outpatient and intensive residential therapy, and transcranial magnetic stimulation. Given the severity and refractoriness of her symptoms, she was enrolled in this clinical trial for invasive brain mapping to guide personalized, multi-site DBS for her OCD.

## Results

### Stimulation-Response Mapping

Twelve stereoelectroencephalography electrodes, each with sixteen contacts, were implanted bilaterally in the orbitofrontal cortex (OFC) [5], anterior cingulate cortex (ACC) [6, 7], dorsal cingulate cortex (DCC), ventral capsule/nucleus accumbens (VC/NAc) [8], ventral capsule/bed nucleus of the stria terminalis (VC/BNST) [9, 10], and anterior medial subthalamic nuclei as well the neighboring zona incerta (amSTN/ZI) [11, 12] (Fig 1A). The post-operative CT was registered to the pre-operative MRI to verify the location of each electrode contact. We then performed extensive stimulation mapping testing (Fig 1B) during a 10-day inpatient monitoring stay. In Phase 1, we conducted safety testing with brief stimulation trains across 29 anatomical regions of interest at currents ranging from 1-6 mA and durations of 1-30 seconds to rule out sites associated with adverse effects, such as involuntary movements, autonomic effects, excessive mood elevation concerning for mania, or spiking activity on iEEG monitoring concerning for progression to a seizure. In Phase 2, we delivered 5-minute stimulation trains at 21 electrode contact pairs to identify sites associated with improvement in self-reported OCD symptoms while again monitoring for adverse events. Symptoms were assessed using visual analogue scales (VAS) for obsessions, compulsions, OCD-related distress, depression, anxiety, and energy. Based upon these results, we identified six promising stimulation sites associated with the largest reductions in OCD (Fig 1B, C). In Phase 3, these top candidate sites were tested using a 20-minute randomized, sham-controlled stimulation paradigm. Both the participant and clinicians were blinded to stimulation site, with repeated trials of stimulation (Fig 1B, D left). A provocation paradigm was used during testing to elicit OCD symptoms if the participant was asymptomatic. During this phase, two targets within the right ventral capsule (VC), above the neighboring the NAc and BNST (Fig 1D), were associated with the most improvement in OCD symptoms (Fig 1D). VAS changes in energy, anxiety, and depression were also observed, though these were small relative to the improvement in OCD symptoms with sham-controlled stimulation.

**Figure 1:**
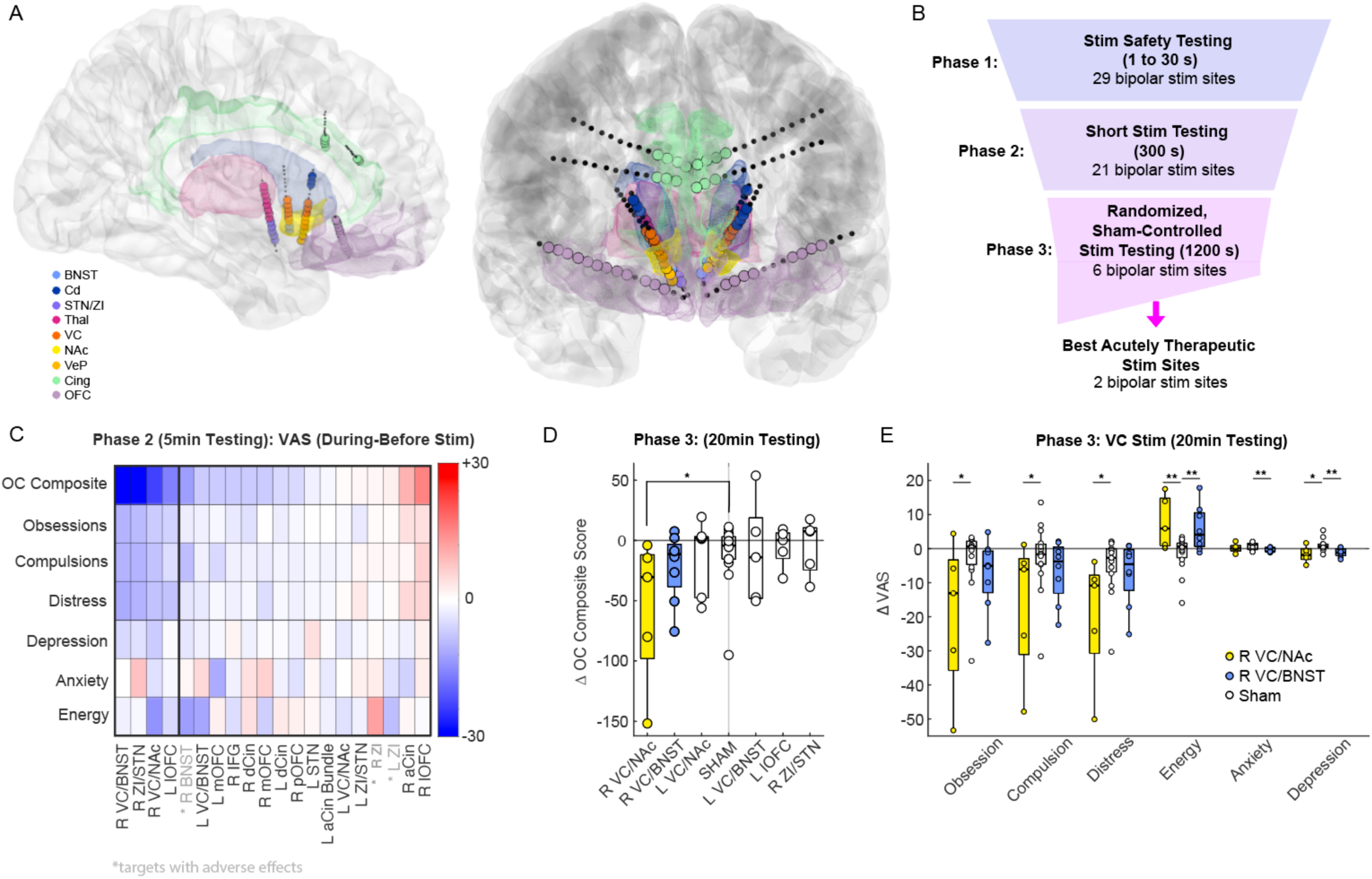
Invasive Brain Mapping to Identify Personalized Neuromodulation Targets. A) Location of anatomically verified intracranial electrodes, B) Stimulation testing paradigm during invasive monitoring, C) Mean change in VAS before and during stimulation across sites during Phase 2 efficacy testing, D) Changes in VAS-OC composite score before and during stimulation across six top candidates sites and with sham during Phase 3, E) Change in VAS subscales before and during R VC/NAc, R VC/BNST, and sham stimulation during Phase 3 (* p<0.05, ** p<0.01, significance based upon permutation test) (left).

### Biomarkers of Symptom State and Target Engagement

To characterize the relationship between CSTC activity and OCD symptoms, we collected intracranial EEG recordings during VAS assessments (Fig 2A). An OCD composite total score representing severity of OCD symptoms was calculated as the sum of VAS scales for obsessions, compulsions, and OCD-related distress, which were all strongly correlated (Extended Data Fig 1). We then identified electrophysiological correlates of the self-report scales and composite total score, focusing in particular on high frequency activity (HFA: 30-95Hz), which is a measure of local neuronal population activity. We found that HFA within the orbitofrontal cortex, anterior/dorsal cingulate, caudate, and ventral thalamus correlated significantly with the OCD total score (Fig 2B). We also found that beta and mid-frequencies throughout the CSTC were correlated with the severity of OCD symptom whereas low-frequency (delta) power at some CSTC sites was anti-correlated with OCD symptoms (Extended Data Fig 2). Together, these data are consistent with the hypothesis that higher activity within a distributed CSTC circuit is associated with greater severity in OCD symptoms.

**Figure 2:**
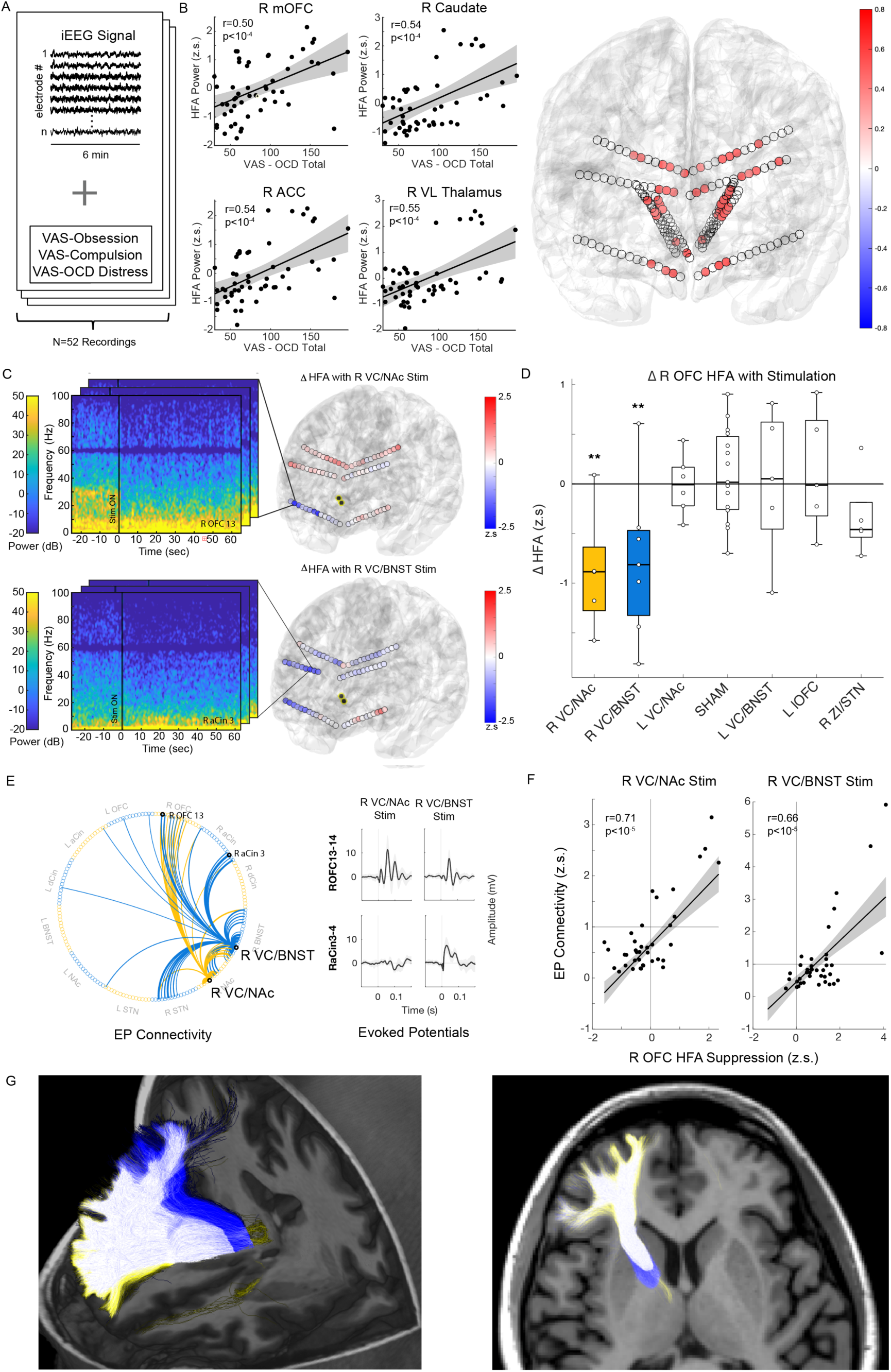
CSTC high frequency activity (HFA) correlates with OCD symptom severity and acute therapeutic stimulation in ventral capsule suppresses orbitofrontal HFA. A) Biomarker development from intracranial recordings and VAS self-report, B) Examples of correlation between HFA and VAS-OCD total scores across representative brain regions within CSTC network (left). Shaded region represents 95% confidence interval. Summary of significant HFA vs VAS-OCD total correlations across implanted electrodes (significance defined by Bonferroni-corrected Permutation Test for 174 recording sites). Color bar represents Pearson correlation. C) Example spectrogram of R lateral orbitofrontal spectral changes with R VC/NAc stimulation (top left). Example spectrogram of R anterior cingulate spectral changes with R VC/BNST stimulation (bottom left). Summary of change in cortical HFA (z-scored to baseline) with stimulation during 20-minute recordings across cortical recording sites (right). Contacts highlighted in yellow represent stimulation contacts. D) Trial-by-trial change in average HFA across right orbitofrontal recording sites with stimulation at candidate targets (** p < 0.01, permutation test comparing active stimulation to sham, 20-minute recordings). E) Directed graph of single-pulse stimulation N1 (10-50 ms post-stimulation) EP responses (greater than 1 standard deviation) from R VC/NAc (yellow) and R VC/BNST (blue) contacts with examples of EP waveforms. F) Correlation of N1 EP magnitude and HFA suppression across cortical electrodes. Line represents linear fit and shaded grey region is 95% confidence interval. G) Tractography derived from the R VC/NAc (yellow), R VC/BNST (blue), or both contacts (white).

We next wanted to characterize the neurophysiological effect of stimulation at the acutely therapeutic R VC/NAc and R VC/BNST sites on this CSTC HFA biomarker of OCD severity. Stimulation at each target was associated with a significant suppression of OFC HFA (Fig 2C, D). In addition, R VC/BNST also led to suppression in the anterior cingulate (Fig 2C). The suppression of OFC activity was specific to stimulation at the R VC/NAc and R VC/BNST sites and not observed with stimulation at other candidate targets (Fig 2D). We then sought to understand the electrophysiological connectivity between the right candidate VC sites and OFC, ACC, and other components of the CSTC network as another measure of target engagement. We measured the N1 component of the evoked potential (EP) response across recording sites in response to single pulse stimulation of the R VC/NAc and R VC/BNST targets (Fig 2E). Both R VC/NAc and R VC/BNST stimulation elicited strong N1 EP responses within the OFC. In addition, the R VC/BNST also had strong EP connectivity to the ACC. These results suggest that both therapeutic VC stimulation sites provide electrophysiological input to the OFC (Fig 2E). Notably, the strength of the EP response correlated with the magnitude of suppression of high frequency activity with stimulation across cortical sites (Fig 2F).

In addition, we used tractography from diffusion-weighted MRI to determine structural connectivity from the therapeutic VC sites. Both sites were located within a component of the anterior thalamic radiation (ATR), which contains tracts connecting anterior thalamus with the OFC (Fig 2G). The VC/BNST stimulation site also had dense connectivity to the ACC similar to the spatial pattern of EP and stimulation-induced suppression.

### Multi-site DBS to Treat OCD

SEEG electrodes were explanted at the end of the invasive monitoring stay. Eight weeks later, the participant underwent four-lead DBS surgery with bilateral Medtronic Percept RC implantable pulse generators. Based on the brain mapping results, the R VC/NAc and R VC/BNST sites were implanted with chronic DBS leads. Leads were also placed within the L VC/NAc, which had shown a modest but less reproducible improvement in OCD symptoms compared to the R VC sites (Fig 1D), and the left ACC was implanted for long-term chronic sensing using the Percept device. Initial DBS programming took place two weeks after DBS surgery. DBS was first delivered bilaterally to VC/NAc, similar to the conventional DBS configuration for OCD, resulting in a rapid but modest improvement in OCD symptoms. The following week, the personalized R VC/BNST DBS was added, leading to a larger improvement in OCD symptoms with combined stimulation (Extended Data Fig 4). One month from initial programming, the participant experienced a 41% improvement in her YBOCS score (YBOCS 20) compared to baseline (YBOCS 34).

## Discussion

These results of this case are consistent with models hypothesizing that elevated CSTC network activity mediates OCD symptoms [13–15]. In this model, therapeutic VC stimulation disrupts basal thalamocortical input, reducing activity in the orbitofrontal and cingulate cortices, leading to reductions in OCD symptoms [16, 17]. Prior studies have demonstrated that stimulation near the ATR projecting to OFC is associated with better DBS outcomes in OCD [18–23]. This model is consistent with theories that VC DBS acts as a reversible informational lesion [24] and the existing clinical evidence that capsulotomy can effectively treat refractory OCD [25, 26]

This case provides the first proof-of-concept that acute stimulation at specific targets can rapidly and reproducibly reduce OCD symptoms during an invasive monitoring stay and be used to guide multi-site DBS. Our observation of OCD symptom reduction within a short timeframe is notable, as prior reports suggest that the ‘wash-in’ time for OCD benefit from DBS typically takes days to weeks [2]. In fact, most intra-operative testing and VC DBS programming assess changes in ‘mood’, ‘energy’, and ‘anxiety’ since changes in core OCD symptoms are not believed to be discernable during such short time frames [27]. Our ability to detect rapid changes in OCD symptoms may be due to several factors. First, the 10-day invasive monitoring period allowed us to test a wide range of stimulation contacts and configurations for longer durations than a standard outpatient visit. This provided a unique opportunity to identify stimulation settings that yielded robust immediate effects on OCD symptoms. Second, we planned symptom provocations during this monitoring period to better control OCD symptoms, disentangling therapeutic benefits specific to OCD from other confounds like mood improvements. Lastly, the randomized sham-controlled testing allowed us to identify therapeutic effects specifically due to the stimulation, as opposed to other confounds such as non-specific placebo effects or natural habituation of symptoms. The ability to extensively test and assess a wide range of stimulation settings to identify optimal parameters during an invasive brain mapping stage to guide the development on chronic DBS opens up the possibility of personalized neuromodulatory therapies.

Further, our invasive brain mapping approach allowed us to identify biomarker of OCD symptom severity and verify that our personalized neuromodulation targets are engaging these circuits. While several studies have implicated CSTC pathophysiology in OCD, this is the first study to directly demonstrate that temporal shifts in the electrophysiological activity across the CSTC network strongly correlate with the symptom severity over time within an individual. Moreover, VC stimulation at both targets induced a strong suppression of orbitofrontal activity were associated with acute symptomatic improvement, suggesting a critical role of OFC in mediating symptoms within the CSTC circuit. It is possible that our novel stimulation paradigm targeting both VC/NAc and VC/BNST within thalamocortical tracts was able to more effectively suppress orbitofrontal activity than could be achieved with stimulating either site alone. This could explain why the combined mutli-site stimulation strategy yielded a greater acute improvement in symptoms than standard bilateral VC/NAc DBS in this subject.

This study has some limitations. First, the stimulation mapping protocol inherently favors stimulation targets that can produce a rapid effect. Given that the inpatient monitoring stay only lasts 10 days and the large number of sites that were tested, we could only trial stimulation for a limited duration of time at each target. It is possible that candidate sites with slower onsets of response, which could still be therapeutically beneficial with chronic stimulation, may be missed in our stimulation testing paradigm. Second, it is unclear whether the biomarkers of OCD symptoms and therapeutic response that we observed during the invasive brain monitoring will generalize to the naturalistic context of the home environment. With DBS devices capable of performing longitudinal intracranial recordings and delivering stimulation, we can determine if biomarkers and stimulation findings from inpatient monitoring will generalize to real-world, long-term treatment [28–31]. We aim to address these questions in subsequent phases of this study.

It remains to be seen whether the personalized stimulation sites and biomarkers identified in this subject will generalize to other participants. If these findings do generalize to other future participants, this could indicate that there may be a common circuit dysfunction underlying OCD that can be consistently targeted to achieve therapeutic responses. This would mean that a limited number of DBS targets could treat OCD without the need for additional personalization [32]. Alternatively, we may find that the optimal stimulation sites and OCD biomarker network are distinct across individuals. This would justify a precision medicine approach, relying on mapping for personalization of therapy. Additional cases should allow us to distinguish between these different possibilities.

## Methods

### Participant

In this n-of-1 study/case study, the patient was a 20s year-old woman with childhood onset severe, treatment refractory OCD (YBOCS 36) unresponsive to at least 3 SSRIs, 1 SNRI, 4 antipsychotic augmentation trials, and augmentation with Clomipramine. She also was not responsive to multiple trials of outpatient and residential intensive therapy for her OCD as well as TMS over the dorsomedial prefrontal cortex/anterior cingulate cortex.

### Surgical procedure

The patient gave written informed consent for participation in a clinical trial of Personalized DBS for OCD Guided by Stereoencephalography Mapping (NCT06347978), approved by the UCSF institutional review board and Food and Drug Administration (FDA). The patient was surgically implanted with twelve stereoelectroencephalography (SEEG) electrodes (PMT Corporation) within the most promising sites bilaterally for modulating OCD based on published literature: VC/NAc [2], VC/BNST [9, 10], and amSTN/ZI [11, 33]. Additionally, we targeted anterior and posterior dorsal sites in the ACC as well as the OFC as these regions are the targets of TMS for OCD [5, 6] and cingulotomies [34]. SEEG leads with 2.5mm center-to-center spacing were used in targeting the subcortical VC/NAc, VC/BNST, and amSTN/ZI, and leads with 3.5mm spacing were used in ACC and OFC. Surgical targeting was planned in Brainlab iPlan Cranial Software using a combination of anatomic and diffusion tractography based targeting. Computed tomography (CT) was used intra- and post-operatively to confirm electrode placement. No complications of surgery occurred. Post-operative CT was registered to pre-operative MRI to determine the anatomic location of each electrode contact. All electrode locations were analyzed by a neuroradiologist (LS), who confirmed that a subset of the 16 contacts on each lead was in the anatomic area of interest. Exploratory intracranial stimulation and recording took place over a 10-day period. At the end of the 10-day hospitalization, the electrodes were explanted.

### Stimulation testing

Briefly, we conducted safety stimulation testing of a preselected set of 49 stimulation targets through a bipolar stimulation survey, using charge-balanced biphasic stimulation with a frequency of 100 Hz and pulse width of 100 μs. Stimulation current amplitudes and durations were progressively increased from 1–6 mA for 1,3, and 30 seconds. After an initial broad survey, we selected a reduced set of 21 sites for 5-minute preliminary stimulation efficacy testing again at progressively higher currents from 2-6 mA. The six stimulation configurations associated with the greatest acute improvement in OCD symptoms were then tested during longer stimulation periods (20 min) with randomized blinded, sham-controlled stimulation in which the assessing clinicians and participant were unaware of the stimulation parameters being tested.

### Clinical measures

We measured symptoms intensity by continuously collecting visual analog scales (VAS) using REDCap electronic data capture tools hosted at UCSF. We collected VAS-scales for: Obsessions, Compulsions, OCD-related Distress, Anxiety, Depression, and Energy. They were measured on a continuous scale from 0 to 100. We found that OCD sub-scores were highly correlated (Extended Data Fig 1). As a consequence, we used a composite total score summing all the scores (obsessions, compulsions, OC-distress) to reduce the dimensionality of the analysis, when necessary.

To select the most promising targets during Phase 2, we collected the mean VAS value during the 5 minutes preceding the start of the stimulation (baseline score) and the mean VAS during the 5 minutes of stimulation. We ranked targets by order of best change in a single recording, i.e. biggest decrease in OC composite score (Fig. 1C). To assess the effect of stimulation on the scores, we collected the mean VAS value during the 5 minutes preceding the start of the stimulation and the mean VAS during the 20 minutes of stimulation, including sham stimulation sessions. We extracted the change in VAS for all trials (N= 5 to 9 trials depending on stimulation target, 15 sham trials). We compared the change due to the stimulation of each target to the change due to sham stimulation using permutation tests with 10,000 permutations (Fig. 2D, E).

### iEEG recording

Intracranial electroencephalogram (iEEG) recordings were collected from implanted SEEG electrodes. Stimulation through these electrodes either as single pulses or continuous stimulation was used to assess connectivity and assess for clinical and network-wide effects. iEEG recordings and evoked potentials mapping were acquired using a 256-channel Nihon Kohden clinical system and secondary data stream at a sampling rate of 10 kHz.

### Biomarker Analysis

iEEG recordings were divided into biomarker sessions, each consisting of 6-minute segments aligned with the time periods immediately preceding each symptom self-report. iEEG timeseries was first re-referenced to a bipolar configuration. Data was then downsampled to a rate of 1024 Hz before undergoing high pass filter above 0.5Hz. A notch filter was then applied to remove line noise artifact at 60Hz and harmonics (120Hz, 180Hz, 240Hz).

Preprocessed data was then analyzed in the time-frequency domain using a Morlet transform. A Morlet wavelet with 6 cycles was applied, using 50 logarithmically spaced frequency filters between 1 and 250 Hz. Time-varying spectral power was then calculated across each of the 50 center frequencies. An automatic artifact rejection procedure was then employed by identifying time segments with broadband power exceeding five standard deviations from the median of the robust z-score distribution. These timepoints were excluded from further analysis. For the remaining clean time segments within each biomarker session, median spectral power was calculated for each center frequency. Frequency band-specific power was then calculated by averaging the median spectral power for all center frequencies falling within the bounds of each canonical frequency band (Delta: 1-4Hz, Theta: 4-8Hz, Alpha: 8-13Hz, Beta: 13-30Hz, HFA: 30-95Hz).

Now having a representation of median band-specific power within each frequency band for each biomarker session (6-minutes directly preceding each self-reported VAS rating), we explored the relationship between each channel’s power across time with fluctuations in the patient’s self-reported OCD symptoms. We did this by calculating the Pearson correlation between each channel’s frequency-specific powers and VAS-Composite score (calculated by summing three VAS-OCD sub-scores). We employed a permutation test to assess the statistical significance of these correlations, permuting the temporal order of VAS-OCD Composite scores 10,000 times. After generating a null distribution of correlation coefficients, the p-value for the observed correlation coefficient was calculated.

### Stimulation Effects Analysis

iEEG recordings were segmented into blocks of time spanning 5 minutes before to 20 minutes after the onset of stimulation and downsampled to 512Hz. Signals were visually inspected to exclude trials exhibiting poor signal. A notch filter was then applied to remove line noise artifact at 60Hz and harmonics (120Hz, 180Hz, 240Hz) and re-referenced to a bipolar montage.

To assess the distributed electrophysiological effect of the stimulation in cortical areas, we first bandpass-filtered the time course of each channel’s signal into an HFA band (30-95Hz), which is known to reflect local activity. The 95Hz upper limit was used to avoid the artifactual effects of the 100Hz stimulation frequency. We calculated for each stimulation segment (time resolution = 5 s, sampling frequency = 512 Hz, frequency resolution = 0.2 Hz; figure 2C, left) (see extended Fig 3). Power spectra were then z-scored to the 30 second baseline prior to stimulation and averaged across the HFA band. The change in HFA before and during stimulation was calculated from the difference in the average power comparing the 30 seconds prior to and 20 minutes after the onset of stimulation across contacts for the R OFC lead for each stimulation trial. For each stimulation target, elicited changes were then compared to changes from sham trials using a permutation test (n = 10000, figure 2D).

### Diffusion Tractography

We acquired presurgical magnetic resonance imaging on a GE 3T Signa Premier with a 32 channel head coil, including a T1-weighted 3D BRAVO (1mm isotropic; repetition time 6.22ms, echo time 2.468ms, inversion time 600ms) and diffusion weighted dual spin echo HARDI sequence (4 b=0 and 83 b=2000s^2^/mm diffusion encoding directions split across a blip-up and blip-down acquisition; FOV 220mm, acquisition matrix 110×110, slice thickness 2mm). Diffusion weighted image noise and Gibbs ringing artefacts were reduced using MRTrix3, followed by correction for spatial distortions and head-motion via TOPUP and Eddy. MRtrix Single-shell Multi-Tissue separated the white matter signal from that of the grey matter and cerebrospinal fluid, identifying single-orientation voxels using the Dhollander method and modelling fiber orientation distributions using spherical harmonics. The T1 weighted image was N4 bias corrected, skull stripped using HD-BET, and its rigid transform calculated to both the post-operative CT and the first pre-operative distortion-corrected b=0 diffusion image. Coordinates for leads in the VC/BNST activating the lateral OFC were identified on a post-operative CT image and transformed into 2mm-radii spheres of presumed tissue activation in diffusion space, via the aforementioned rigid registrations. Tractography using iFOD2 generated 50,000 streamlines unidirectionally seeded from these regions of presumed tissue activation, that passed through a coronal plane in the frontal lobe ∼7mm anterior to these spheres and terminated according to default criteria. For confirmation, we also performed tractography in the reverse direction, from the lateral OFC to these lead spheres, expecting that, should this connection be implausible, processing would take an extremely long time to complete and/or result in a high proportion of aberrant streamlines.

### Evoked Potentials Connectivity

Briefly, single-pulse evoked potential (EP) is a functional connectivity measure that describes the directional influence of one brain region on another. Intracranial EEG and intracortical stimulation offer a rare opportunity to probe directional connectivity through the delivery of a single brief stimulation pulse and examination of the evoked potentials at other sites. Evoked potential mapping was performed on day 1. We delivered bipolar single pulse stimulation at 3mA and 1 Hz for 20 s to adjacent contact pairs in each brain region.

We quantified the evoked potential responses to single pulse stimulation at all other contacts by *z*-scoring voltage against 50 ms of pre-stimulus baseline per trial, calculating the mean z-scored evoked potential across 20 trials by measuring the root mean square power of the averaged evoked potential within N1 (10–50 ms) time window. We then applied a threshold on evoked potentials with a magnitude greater than one to generate a directed network graph.

### Provocation Paradigm

Prior to and during the invasive brain monitoring stay, the participant worked with a psychologist and the team psychiatrist to develop a hierarchy of provocative exposures based upon the participant’s harm-based obsessions to elicit OCD symptoms. If the participant’s OCD symptoms declined during stimulation testing, a provocation was performed to increase symptoms to a clinical range of severity that allowed stimulation settings to be trialed so a therapeutic benefit could be observed.

### DBS Surgery and Programming

Medtronic Sensight DBS leads (B33015) were implanted in the right VC/NAc, right VC/BNST, left VC/NAc, and left ACC and connected to bilateral Percept RC IPGs. Right leads were connected to the right IPG, and the left leads were connected to the IPG. Two weeks later, initial DBS programming was performed. Stimulation was delivered continuously at 130Hz between 7-9mA.

## Acknowledgements

This work was supported by a National Institutes of Mental Health (NIMH) K23MH125018 (A.M.L.), NIMH R21MH130914 (A.M.L), Foundation for OCD Research (A.M.L.), Foundation for the NIH (A.M.L), and NARSAD Young Investigator grant from the Brain & Behavior Research Foundation (A.M.L.).

## Contributions

A.M.L., E.F.C, A.D.K., initiated the work and supervised the study. K.K.S. and P.A.S. contributed to the conceptualization and evaluation of the study protocol. A.K., J.A, K.K.S., L.P.S, J.M.F, A.B.A, C.A.R, A.T.M, and N.B. helped with data collection. A.K., J.A., A.N.I, L.P.S, L.B.R, K.K., analyzed the data. A.M.L drafted the manuscript. A.D.K. and E.F.C. finalized the manuscript. All authors approved the work and take responsibility for its integrity.

## Author information

### Contributions

AML

AK

KKS

ANK

LPS

LR

KK

JMF

ABA

TN

DA

ATM

NB

AA

PAS

EFC

ADK

## Data Availability

The data used to produce the results and figures in this paper are available at request.

---

**Extended Data Fig 1:**
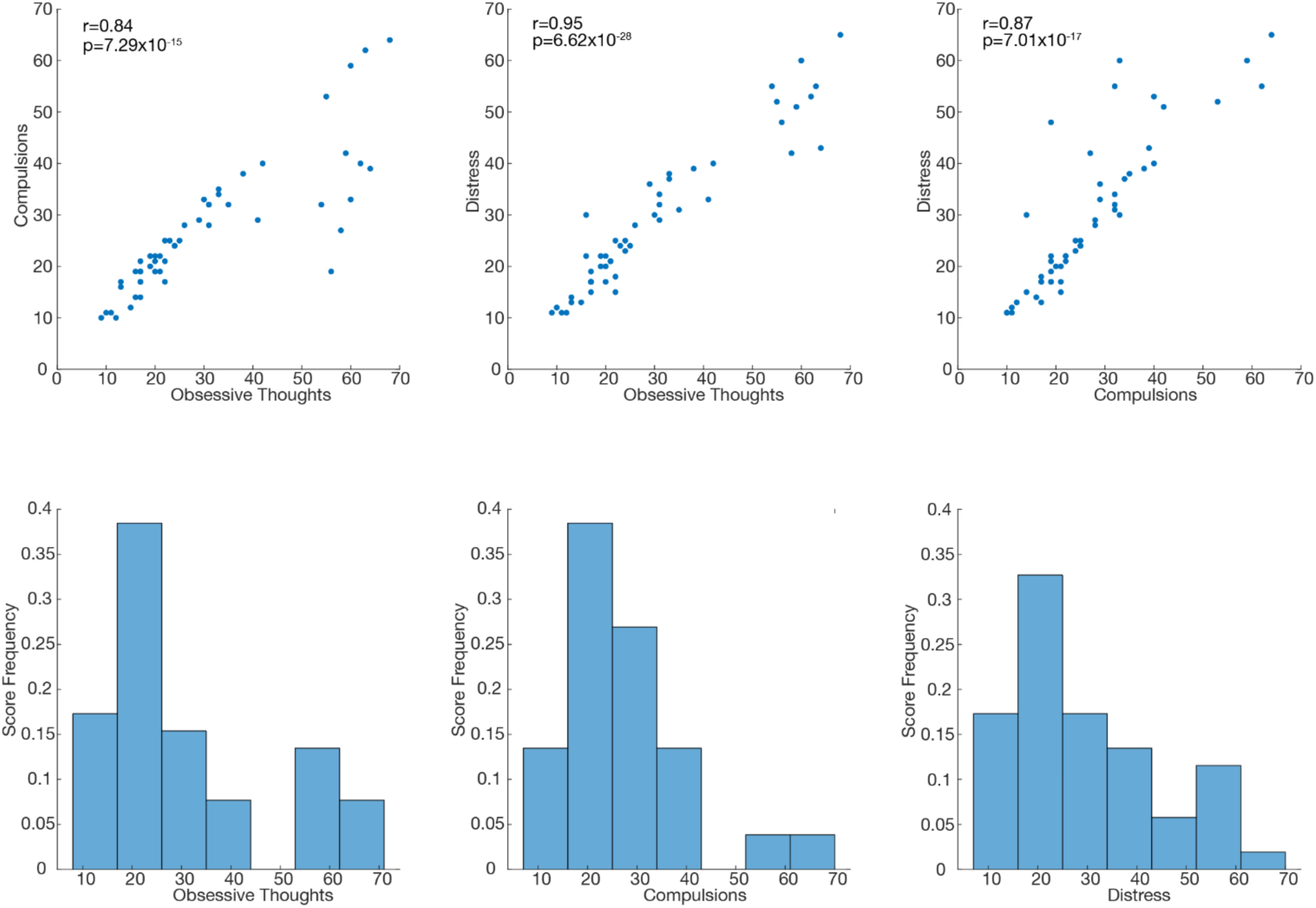
VAS OCD Subscales. Correlation between OCD subscale scores (top). Histograms of OCD subscale scores (bottom)

**Extended Data Fig 2:**
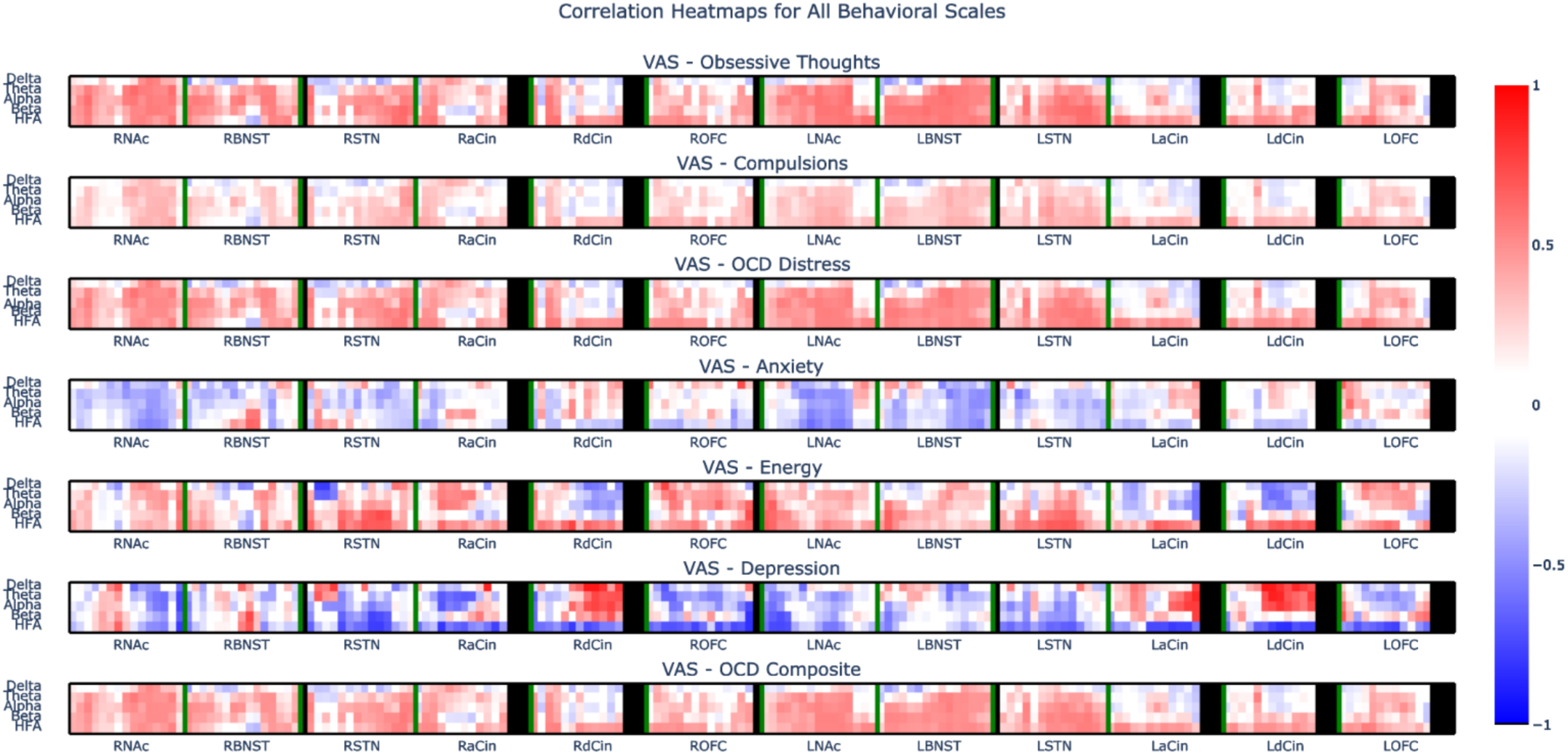
Correlation between VAS scores and power across recording sites. Heatmap of correlation between visual analogue scales for obsessive thoughts, compulsions, OCD distress, anxiety, energy, depression, and the OCD total composite score and power in the delta, theta, alpha, beta, and HFA (30-95-Hz) bands. Red indicates positive correlation. Blue indicates anti-correlation.

**Extended Data Fig 3:**
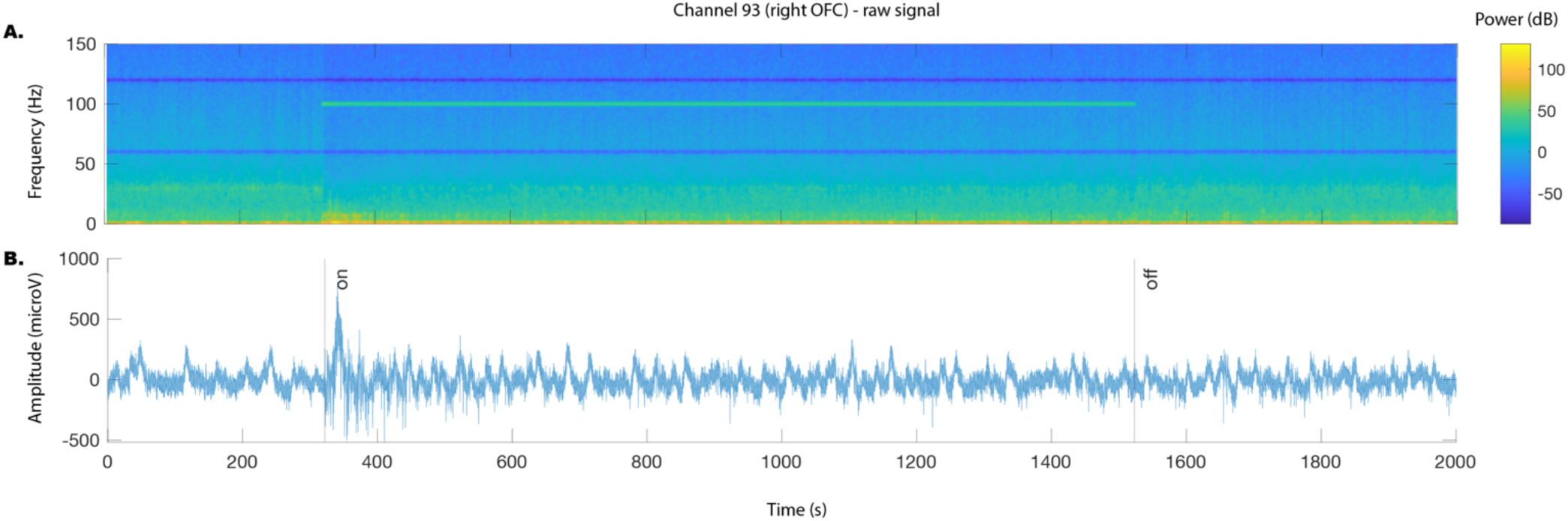
Example recording of an RVC/NAc stimulation trial. Example A) spectrogram and B) LFP recording for right lateral OFC recording (same electrode as in Fig 2C) during 100Hz stimulation. Preprocessing for LFP recordings includes notch filter at 60 and 120 Hz, removal of high amplitude artifacts. Black lines indicate stimulation start and stop.

**Extended Data Fig 4:**
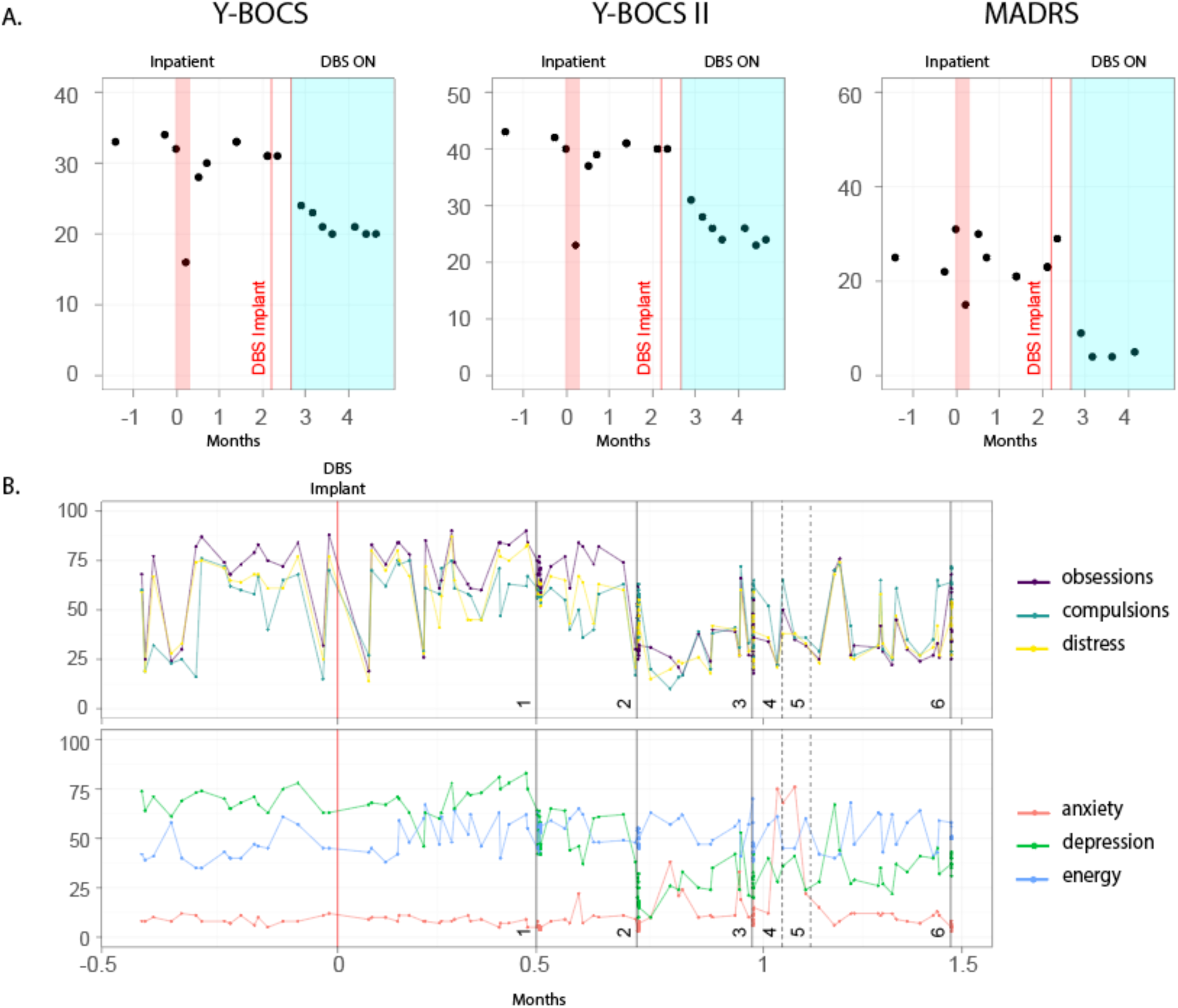
Response to DBS. A. Changes in clinical scores over time, before, during and after in -patient stay. B. Changes in self-reported visual analog scales before and after DBS implant and changes in DBS settings. The black lines show the dates of programming sessions; solid lines are in-office DBS changes that were done by a clinician whereas dashed lines show changes in programming initiated b the participant. For session 1, stimulated targets were R VC/NAc and L VC/NAc only. Starting session 2, R VC/BNST stimulation was added to R VC/NAc and L VC/NAc. Amplitudes levels were the same across stimulation sites: (session1) 7mA; (2) 7mA; (3) 9mA; (4) self-changed to 7mA; (5) self-changed to 9mA; (6) 9mA.

